# Health indicators as a measure of individual health status: public perspectives

**DOI:** 10.1101/2022.03.04.22271701

**Authors:** Temiloluwa Sokoya, Yuchun Zhou, Sebastian Diaz, Timothy Law, Lina Himawan, Francisca Lekey, Lu Shi, Ronald W. Gimbel, Xia Jing

**Author notes:** Corresponding Author Xia Jing, MD, PhD, Edwards Hall 511, Department of Public Health Sciences, College of of Behavioral, Social, and Health Sciences, Clemson University, Clemson, SC 29634.

## Abstract

**Objective:** We examined the perspectives of the general public on 29 health indicators to provide evidence for further prioritizing the indicators, which were obtained from the literature review. Health status is different from disease status, which can refer to different stages of cancer.

**Design:** This study uses a cross-sectional design.

**Setting:** An online survey was administered through Ohio University, ResearchMatch, and Clemson University.

**Participants:** Participants included the general public who are 18 years or older. A total of 1153 valid responses were included in the analysis.

**Primary outcomes measures:** Participants rated the importance of the 29 health indicators. The data were aggregated, cleaned, and analyzed in three ways: (1) to determine the agreement among the three samples on the importance of each indicator (IV = the three samples, DV = individual survey responses); (2) to examine the mean differences between the retained indicators with agreement across the three samples (IV = the identified indicators, DV = individual survey responses); and (3) to rank the groups of indicators after grouping the indicators with no mean differences (IV = the groups of indicators, DV = individual survey responses).

**Results:** The descriptive statistics indicate that the top-five rated indicators are drug or substance abuse, smoking or tobacco use, alcohol abuse, major depression, diet and nutrition. The importance of 13 of the 29 health indicators was agreed upon among the three samples. The 13 indicators were categorized into seven groups. Groups 1-3 were rated as significantly higher than Groups 4-7.

**Conclusions:** This study provides a baseline for prioritizing further the 29 health indicators, which can be used by electronic health records or personal health record system developers. Currently, self-rated health status is used predominantly. Our study provides a foundation to track and measure preventive services more accurately and to develop an individual health status index.

**Strengths and limitations of this study:**

- The work establishes the foundation to measure individual health status more comprehensively and objectively
- The work reflects perspectives from three communities with a relatively large sample size
- The work provides the foundation to prioritize the 29 health indicators further
- With real-world longitudinal data, the public perspective data on individual health status measurement would be verified and validated further

## Introduction

Disease status, such as cancer stage, has been used in routine clinical practice to determine more accurate treatment plans. Health-related indicators, such as mortality, morbidity, and life expectancy for the population group, also have been used. Few studies, however, focus on more comprehensive and objective measures of individual health status. Self-rated health status has been identified as a reliable indicator for an individual’s overall health status[1, 2], but it is subjective, and the rating criteria are unclear. Although there has been research on health indicators used for the measurement of care quality[3] as well as social and behavioral measures in electronic health record (EHR) systems [4, 5]; more comprehensive, objective health indicators of an individual’s health status, which can be used to measure health status more accurately, objectively, and consistently as well as to determine preventive medicine services and their outcomes[1, 6], are lacking. When a healthcare paradigm shifts from treatment to prevention[7, 8], the accurate, objective, and convenient measurement of preventive services and the long-term outcomes of such services become an urgent need.

Individual health status refers to a person’s overall physical, mental, and social well-being as well as freedom from illness or injury, whereas individual disease status refers to a person’s physical or mental symptoms with or without diagnosis[9]. Accurate individual health status measures can be utilized to guide customized preventive services and lifestyle suggestions as well as being applied to population health. This can be accomplished by aggregating an individual’s health data into meaningful groups. Chronic diseases are increasingly costly, and most can be prevented or delayed via preventive services, which need to be provided in a routine and consistent manner[7, 8] with the potential to control healthcare costs.

The Institute of Medicine (IOM) reviewed measures of social and behavioral domains, as seen in EHR[4, 5]. The identified 17 domains and their measures provided a foundation for the Office of National Coordinator’s (ONC) EHR Meaningful Use reporting requirements[4, 5]. In 2015, the Centers for Disease Control and Prevention’s National Center for Health Statistics released 15 selected health indicators based on data from the National Health Interview Survey [10]. Other research[1, 2, 6, 11] also considered health indicators; however, none focused on more comprehensive, objective measures on an individual’s health status. Although preventive medicine has been recognized for its critical role in health care, such services are not provided consistently to the majority of the population[12]. Because chronic diseases represent a large portion of healthcare expenditures, it is critical to delay or prevent chronic diseases via preventive services[13]. The tracking of health indicators has been reported to help policy-makers note changes needed in coverage and influence policy[14]. Such tracking also helps governments to increase the resources allocated for health[14]. Nevertheless, accurate measurements of preventive services are lacking.

We conducted a literature review on health indicators and determined there are 29 health indicators that can be utilized to measure individual health. We then examined four commercial EHR systems in rural primary care ambulatory settings to explore the availability and presentation of these indicators and found that none of the systems captures ***all*** the indicators[9].

The purpose of the current study is to examine public perspectives on the importance of 29 health indicators to provide a means to prioritize these health indicators, for example, to separate the health indicators into core and secondary sets that can be incorporated into EHR or similar systems[15]. Such health indicators can capture an individual’s health status, thus informing preventive services to make them more accurate, consistent, and convenient without overburdening providers’ data collection workload. These public perspectives can also provide a foundation for the development of an individual health index, which can be used to stratify healthy populations into subgroups based on the corresponding study requirements.

## Methods

### Data collection

An online survey (Appendix A) was administered through Ohio University (Summer 2017), ResearchMatch[16] (Summer 2018), and Clemson University (Summer 2020), i.e., three samples. The study was approved by the Institutional Review Boards at Ohio University (17-X-142) and Clemson University (IRB2019-441). The inclusion criterion for participation in the survey was being 18 years or older. The survey link can be shared by participants. All respondents acknowledged informed consent.

The survey included seven demographic questions and rating items in regard to the importance of the 29 health indicators, i.e., alcohol abuse, body mass index (BMI), diet and nutrition, drug or substance abuse, family history of cancer, physical inactivity, smoking or tobacco use, sun protection, immunization/vaccination, insurance coverage, personal care needs, cancer screening detection, hypertension screening, HIV testing, self-rated health status, blood sugar level, blood triglycerides, HDL cholesterol, LDL cholesterol, total cholesterol, high school diploma, air quality index > 100, supply of dentists, engagement in life, health literacy rate, major depression, having a sense of purpose in one’s life, race and ethnicity, and being unemployed. Definitions of these health indicators are provided within the survey (Appendix B).

After removing invalid data, the final sample yielded 362 responses at Ohio University, 694 at ResearchMatch, and 97 at Clemson University (Appendix C). Items were rated on a scale of 0-10 in the survey used by Ohio University and Clemson University, whereas items in the ResearchMatch sample were measured using a scale of 0-100. Therefore, in the data cleaning process, the data from ResearchMatch were converted to a scale of 0-10 (Appendix D contains the codebook). In the Ohio University survey, there were five health indicators, i.e., blood sugar, blood triglycerides, HDL, LDL, and total cholesterol, for which a scale of 0-11, instead of 0-10, was used. Due to this error, data for these five indicators were removed from the Ohio University dataset. As a result, the total sample size of these five indicators was 791, whereas the total sample size of the other indicators was 1153 (Table 1).

**Table 1.** Descriptive statistics for all 29 health indicators

| Health indicators | ResearchMatch |  |  | Ohio University |  |  | Clemson |  |  | Total |  |  |
| --- | --- | --- | --- | --- | --- | --- | --- | --- | --- | --- | --- | --- |
|  | Mean | Std. Deviation | *n* | Mean | Std. Deviation | *n* | Mean | Std. Deviation | *n* | Mean | Std. Deviation | *n* |
| Drug or substance abuse | 8.75 | 1.5 | 694 | 8.13 | 1.96 | 362 | 8.36 | 1.87 | 97 | 8.53 | 1.71 | 1153 |
| Smoking, tobacco use | 8.8 | 1.52 | 694 | 8.02 | 2.06 | 362 | 8.18 | 1.84 | 97 | 8.5 | 1.77 | 1153 |
| Alcohol abuse | 8.34 | 1.71 | 694 | 7.56 | 2.03 | 362 | 8.06 | 1.64 | 97 | 8.07 | 1.84 | 1153 |
| Major depression | 8.1 | 1.6 | 685 | 7.79 | 1.94 | 362 | 8.03 | 1.57 | 97 | 7.99 | 1.72 | 1144 |
| Diet and nutrition | 8.01 | 1.58 | 694 | 7.8 | 1.93 | 362 | 8.36 | 1.65 | 97 | 7.97 | 1.71 | 1153 |
| Blood sugar level | 7.76 | 1.63 | 694 |  |  |  | 7.59 | 1.75 | 97 | 7.74 | 1.65 | 791 |
| Physical inactivity | 7.9 | 1.68 | 694 | 7.41 | 2.13 | 362 | 7.68 | 1.77 | 97 | 7.73 | 1.85 | 1153 |
| Immunization/vaccination | 7.49 | 2.12 | 694 | 7.67 | 2.3 | 362 | 7.72 | 2.4 | 97 | 7.57 | 2.2 | 1153 |
| Hypertension screening | 7.59 | 1.91 | 694 | 7.17 | 2.29 | 362 | 7.42 | 2.03 | 97 | 7.45 | 2.05 | 1153 |
| LDL cholesterol | 7.43 | 1.85 | 694 |  |  |  | 7.56 | 1.91 | 97 | 7.45 | 1.86 | 791 |
| Blood triglycerides | 7.32 | 1.78 | 694 |  |  |  | 7.34 | 1.95 | 97 | 7.32 | 1.80 | 791 |
| HDL cholesterol | 7.31 | 1.83 | 694 |  |  |  | 7.43 | 1.91 | 97 | 7.32 | 1.84 | 791 |
| Having a sense of purpose in one's life | 7.59 | 1.94 | 685 | 6.67 | 2.53 | 362 | 7.88 | 1.93 | 97 | 7.32 | 2.19 | 1144 |
| Cancer screening detection | 7.22 | 2.06 | 694 | 7.26 | 2.3 | 362 | 7.49 | 2.09 | 97 | 7.25 | 2.14 | 1153 |
| Total cholesterol | 7.2 | 2.02 | 694 |  |  |  | 7.6 | 1.85 | 97 | 7.25 | 2.00 | 791 |
| Health literacy rate | 6.99 | 2.02 | 685 | 7.06 | 2.26 | 362 | 7.34 | 2.01 | 97 | 7.04 | 2.10 | 1144 |
| Personal care needs | 6.82 | 2.08 | 694 | 7.01 | 2.3 | 362 | 7.21 | 2.1 | 97 | 6.91 | 2.16 | 1153 |
| Air quality index > 100 | 6.74 | 1.92 | 685 | 6.76 | 2.13 | 362 | 6.89 | 1.93 | 97 | 6.76 | 1.99 | 1144 |
| Family history of cancer | 6.98 | 2.06 | 694 | 6.37 | 2.24 | 362 | 6.25 | 1.98 | 97 | 6.73 | 2.13 | 1153 |
| Self-rated health status | 6.63 | 2.2 | 694 | 6.62 | 2.15 | 362 | 6.92 | 1.89 | 97 | 6.65 | 2.16 | 1153 |
| HIV testing | 6.62 | 2.36 | 694 | 6.62 | 2.64 | 362 | 6.84 | 2.37 | 97 | 6.64 | 2.45 | 1153 |
| Insurance coverage | 6.4 | 2.88 | 694 | 6.79 | 2.91 | 362 | 7.26 | 2.51 | 97 | 6.6 | 2.87 | 1153 |
| Body mass index (BMI) | 6.86 | 2.28 | 694 | 5.8 | 2.54 | 362 | 6.64 | 2.45 | 97 | 6.51 | 2.42 | 1153 |
| Supply of dentists | 6.53 | 2.02 | 685 | 6.34 | 2.26 | 362 | 6.04 | 1.99 | 97 | 6.43 | 2.10 | 1144 |
| Sun protection | 6.63 | 2 | 694 | 5.73 | 2.3 | 362 | 5.54 | 2.18 | 97 | 6.25 | 2.16 | 1153 |
| Unemployed individual | 6.07 | 2.34 | 685 | 5.52 | 2.68 | 362 | 6.2 | 2.69 | 97 | 5.91 | 2.49 | 1144 |
| Engagement in life | 6.38 | 2.18 | 685 | 4.82 | 2.87 | 362 | 6.4 | 2.33 | 97 | 5.89 | 2.54 | 1144 |
| High school diploma as a health indicator | 5.02 | 2.57 | 694 | 6.04 | 3.07 | 362 | 5.56 | 2.75 | 97 | 5.38 | 2.79 | 1153 |
| Race and ethnicity | 5.28 | 2.53 | 685 | 4.32 | 2.76 | 362 | 5.02 | 2.85 | 97 | 4.96 | 2.67 | 1144 |

The internal reliability of the survey instrument (Cronbach’s alpha) for the overall set and the three subsets related to institutions were calculated.

### Patient and Public Involvement statement: Not applicable

#### Data analytic strategies

Data analyses included rating the 29 health indicators based on their importance. After aggregating data from the three samples, we then conducted a three-step analysis. The first step of the analysis was to determine whether the three samples had a unanimous agreement on the importance of each indicator. We ran a one-way ANOVA with a post hoc test in SPSS v.27 for each indicator to examine any group mean difference (IV = the three samples, DV = individual survey responses). The indicators with no group mean differences across samples were retained for the next step of the analysis.

The second step of the analysis was to examine the mean differences between the retained indicators via a one-way ANOVA (IV = the identified indicators, DV = individual survey responses). Any indicators with no significant mean differences were grouped together because they cannot be ranked.

The third step of the analysis was to rank the groups of indicators after grouping the indicators with no mean differences. We ran a one-way ANOVA with a post hoc test to examine the mean differences between the groups of indicators (IV = the groups of indicators, DV = individual survey responses.) Any significant mean difference between two groups of indicators indicates the ranking order of the two groups.

## Results

Descriptive statistics for 29 health indicators are reported in Table 1, and the descriptive statistics for the demographic information for all respondents are reported in Appendix E. A Levene test was run to test the homogeneity of variance for each indicator before running an ANOVA. This resulted in nine health indicators with homogenous variance (Appendix F) and 20 health indicators with heterogeneous variance (Appendix G). For the nine indicators with homogeneous variance, a one-way ANOVA was used. For the 20 indicators with heterogeneous variance, a one-way ANOVA Welch test was used. As a result, 13 indicators were found to have no significant mean differences among the three samples, which indicates that the survey participants agreed on the importance of these indicators (Table 2). These 13 indicators were retained for the next step of the analysis. There were significant mean differences among the other 16 indicators, which indicates that survey participants disagreed on their importance (Appendix H contains the post hoc results).

**Table 2.**
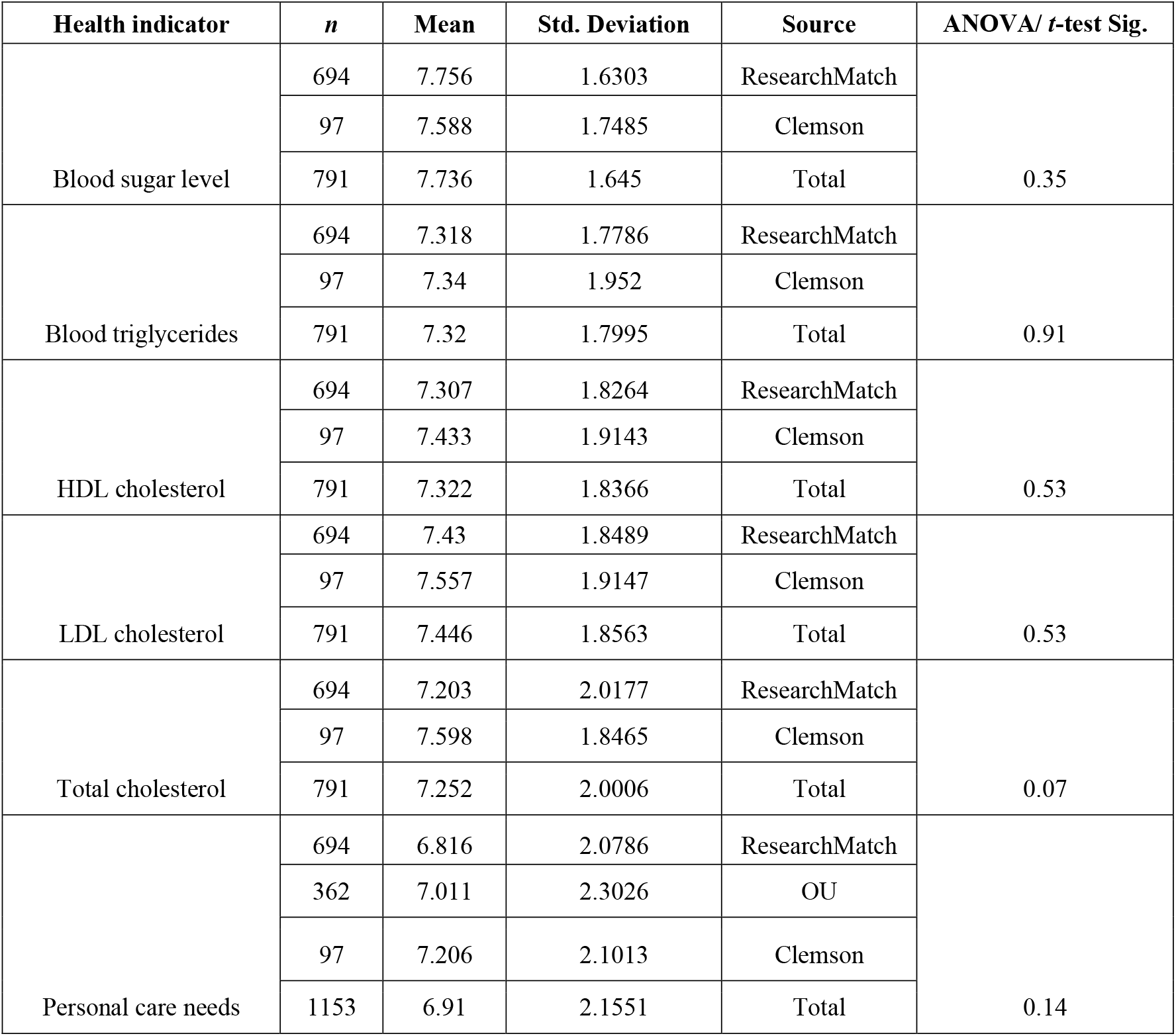
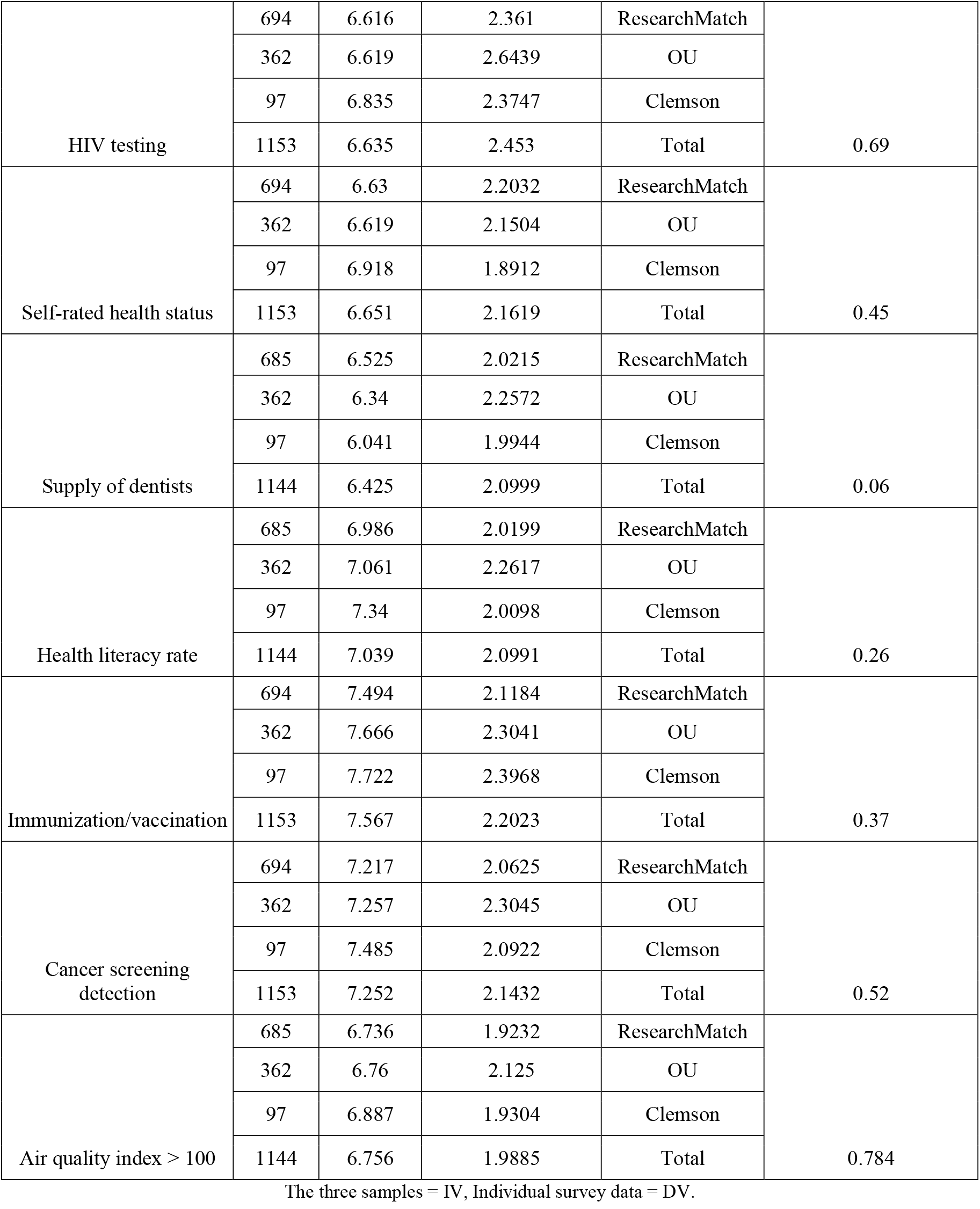
The 13 indicators with non-significant mean differences across the three samples

| Health indicator | *n* | Mean | Std. Deviation | Source | ANOVA/ *t*-test Sig. |
| --- | --- | --- | --- | --- | --- |
| Blood sugar level | 694 | 7.756 | 1.6303 | ResearchMatch | 0.35 |
|  | 97 | 7.588 | 1.7485 | Clemson |  |
|  | 791 | 7.736 | 1.645 | Total |  |
| Blood triglycerides | 694 | 7.318 | 1.7786 | ResearchMatch | 0.91 |
|  | 97 | 7.34 | 1.952 | Clemson |  |
|  | 791 | 7.32 | 1.7995 | Total |  |
| HDL cholesterol | 694 | 7.307 | 1.8264 | ResearchMatch | 0.53 |
|  | 97 | 7.433 | 1.9143 | Clemson |  |
|  | 791 | 7.322 | 1.8366 | Total |  |
| LDL cholesterol | 694 | 7.43 | 1.8489 | ResearchMatch | 0.53 |
|  | 97 | 7.557 | 1.9147 | Clemson |  |
|  | 791 | 7.446 | 1.8563 | Total |  |
| Total cholesterol | 694 | 7.203 | 2.0177 | ResearchMatch | 0.07 |
|  | 97 | 7.598 | 1.8465 | Clemson |  |
|  | 791 | 7.252 | 2.0006 | Total |  |
| Personal care needs | 694 | 6.816 | 2.0786 | ResearchMatch | 0.14 |
|  | 362 | 7.011 | 2.3026 | OU |  |
|  | 97 | 7.206 | 2.1013 | Clemson |  |
|  | 1153 | 6.91 | 2.1551 | Total |  |
| HIV testing | 694 | 6.616 | 2.361 | ResearchMatch | 0.69 |
|  | 362 | 6.619 | 2.6439 | OU |  |
|  | 97 | 6.835 | 2.3747 | Clemson |  |
|  | 1153 | 6.635 | 2.453 | Total |  |
| Self-rated health status | 694 | 6.63 | 2.2032 | ResearchMatch | 0.45 |
|  | 362 | 6.619 | 2.1504 | OU |  |
|  | 97 | 6.918 | 1.8912 | Clemson |  |
|  | 1153 | 6.651 | 2.1619 | Total |  |
| Supply of dentists | 685 | 6.525 | 2.0215 | ResearchMatch | 0.06 |
|  | 362 | 6.34 | 2.2572 | OU |  |
|  | 97 | 6.041 | 1.9944 | Clemson |  |
|  | 1144 | 6.425 | 2.0999 | Total |  |
| Health literacy rate | 685 | 6.986 | 2.0199 | ResearchMatch | 0.26 |
|  | 362 | 7.061 | 2.2617 | OU |  |
|  | 97 | 7.34 | 2.0098 | Clemson |  |
|  | 1144 | 7.039 | 2.0991 | Total |  |
| Immunization/vaccination | 694 | 7.494 | 2.1184 | ResearchMatch | 0.37 |
|  | 362 | 7.666 | 2.3041 | OU |  |
|  | 97 | 7.722 | 2.3968 | Clemson |  |
|  | 1153 | 7.567 | 2.2023 | Total |  |
| Cancer screening detection | 694 | 7.217 | 2.0625 | ResearchMatch | 0.52 |
|  | 362 | 7.257 | 2.3045 | OU |  |
|  | 97 | 7.485 | 2.0922 | Clemson |  |
|  | 1153 | 7.252 | 2.1432 | Total |  |
| Air quality index > 100 | 685 | 6.736 | 1.9232 | ResearchMatch | 0.784 |
|  | 362 | 6.76 | 2.125 | OU |  |
|  | 97 | 6.887 | 1.9304 | Clemson |  |
|  | 1144 | 6.756 | 1.9885 | Total |  |

The three samples = IV, Individual survey data = DV.

In the second step of the analysis, a one-way ANOVA was run for the 13 retained indicators (IV = 13 indicators; DV = individual survey responses). The indicators with no mean differences were grouped together (Table 3) because they were equally important and could not be ranked within a group. As a result, seven groups were formed (Table 3).

**Table 3.**
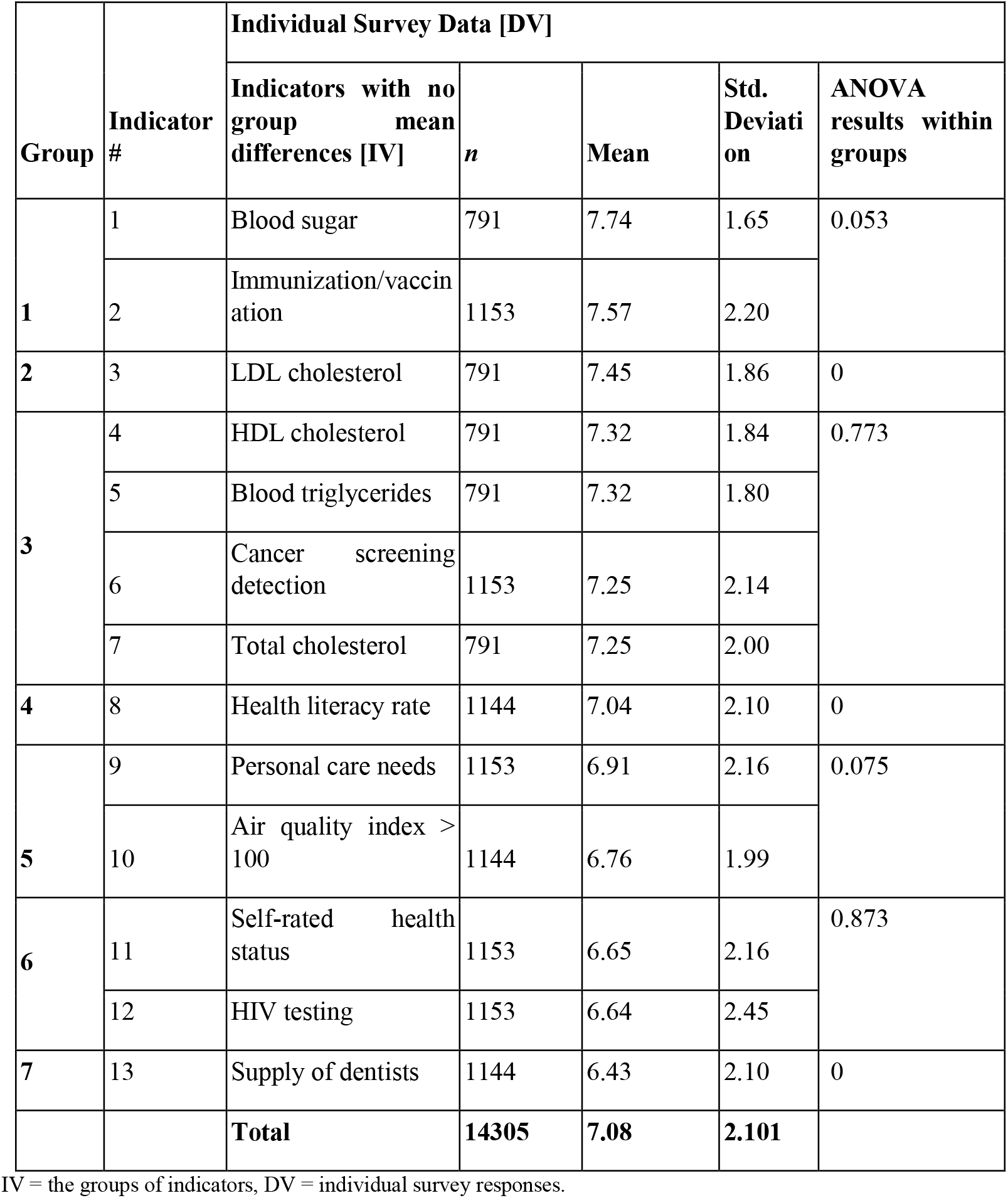
The 7 groups of health indicators with no significant mean differences within groups

In the third step of the analysis, a one-way ANOVA was run for the seven groups of indicators for the purpose of ranking (IV = 7 groups; DV = individual survey responses). There were seven indicators in Group 1, Group 2, and Group 3 (blood sugar, immunization/vaccination, LDL cholesterol, HDL cholesterol, blood triglycerides, cancer screening detection, and total cholesterol). These indicators were found to be significantly more important than the six indicators in Group 4, Group 5, Group 6, and Group 7 (health literacy rate, personal care needs, air quality index > 100, self-rated health status, HIV testing, and supply of dentists; Table 4).

**Table 4.** ANOVA post hoc test results for the seven groups of indicators

| Health Indicator | Health indicator | Sig. |
| --- | --- | --- |
| Group 1 | Group 2 | 0.582 |
| Group 1 | Group 3 | <.001 |
| Group 1 | Group 6 | <.001 |
| Group 1 | Group 7 | <.001 |
| Group 2 | Group 4 | 0.006 |
| Group 2 | Group 3 | 0.678 |
| Group 2 | Group 5 | <.001 |
| Group 2 | Group 7 | <.001 |
| Group 3 | Group 4 | 0.063 |
| Group 3 | Group 5 | <.001 |
| Group 4 | Group 5 | 0.274 |
| Group 4 | Group 1 | <.001 |
| Group 4 | Group 6 | <.001 |
| Group 5 | Group 6 | 0.136 |
| Group 5 | Group 1 | <.001 |
| Group 5 | Group 7 | <.001 |
| Group 6 | Group 7 | 0.207 |
| Group 6 | Group 2 | <.001 |
| Group 6 | Group 3 | <.001 |
| Group 7 | Group 2 | <.001 |
| Group 7 | Group 3 | <.001 |
| Group 7 | Group 4 | <.001 |

IV = the groups of indicators, DV = individual survey responses.

Moreover, among the more important indicators, the two indicators in Group 1 (blood sugar and immunization/vaccination) were found to be significantly more important than the four indicators in Group 3 (HDL cholesterol, blood triglycerides, cancer screening detection, and total cholesterol).

Further, among the less-important indicators, the indicator in Group 4 (health literacy rate) was found to be significantly more important than the two indicators in Group 6 (self-rated health status and HIV testing) and the indicator in Group 7 (supply of dentists). The two indicators in Group 5 (air quality index > 100 and personal care needs) were significantly more important than the indicator in Group 7 (supply of dentists).

The instrument’s (n=29) items showed favorable levels of internal reliability (Cronbach’s Alpha = 0.912), as did each of the three subsets related to institutions where the survey was administered (see Table 5).

**Table 5.** Cronbach’s alpha for the survey instrument (entire survey and subscales)

| Survey components | Data analyzed | Cronbach's Alpha |
| --- | --- | --- |
| **Entire survey** (all items) | All 3 samples | 0.912 |
|  | ResearchMatch | 0.922 |
|  | Ohio University | 0.893 |
|  | Clemson University | 0.925 |
| **Survey subscales** | Health risks and behaviors indicators | 0.795 |
|  | Healthcare | 0.613 |
|  | Healthcare provider supply | 0.831 |
|  | Blood tests in physical exams | 0.934 |
|  | Other health indicators | 0.823 |

## Discussion

### Interpretation of the results

Descriptive analyses reveal drug and substance abuse, smoking and tobacco use, alcohol abuse, major depression, and diet and nutrition are the five most important health indicators, as rated by the study participants. In addition, race and ethnicity, high school diploma, engagement in life, unemployment, and sun protection are the five least important health indicators. Self-rated health status, the most commonly used health indicator that measures an individual’s health status, is ranked in the 20^th^ position.

Among all three samples, the ranking of importance of the 13 health indicators showed agreement (Table 3). Given descriptive statistics and inferential test results, our findings can reasonably be generalized to a broader population beyond our survey respondents. These health indicators, however, are not necessarily more important than the other 16 health indicators. When we compare the 13 health indicators (Table 3) and their corresponding ranks in Table 1, we notice that the 13 health indicators are placed between the 6^th^ and 24^th^ rankings in Table 1. This indicates that there is more agreement among participants in regard to the middle-ranked health indicators than the higher- or lower-ranked ones, i.e., the perspectives are more heterogeneous for those higher- or lower-ranked health indicators.

Among these 13 health indicators found in the seven groups, all groups are not significantly different from their immediate group (Table 4), i.e., there is no significant difference between Groups 1 and 2 (i.e., between n and n + 1). There are, however, significant differences between Groups 1 and 3 to 7 (i.e., between n and any group > n + 1). These results pertain to the further prioritization of health indicators.

Cronbach’s alpha is one of the more cited statistics for informing internal consistency for the items of an instrument. In general, if the Cronbach’s alpha is greater than 0.7, the instrument is considered reliable. The Cronbach’s alpha for the entire survey among three samples is between 0.893 and 0.925, which indicates that we developed a reliable survey instrument. When we look at the subscales, only the healthcare category, which includes vaccination/immunization, insurance coverage, and personal care need, is below 0.7. The items in this category are among the most discussed topics in health care in the United States, it is understandable that the reliability is lower, as the respondents have less consistent perspectives for these items.

### Significance and comparison with related research

Our work provides a more comprehensive understanding of an individual’s health status, particularly as compared to self-rated health status, the most commonly used health status measurement[2]. Although advantages exist for utilizing a single health indicator during clinical encounters, we believe multidimensional measurement of individual health status may be more objective and can provide additional insight for individual health status, particularly if we are concerned with preventive services. To obtain public perspectives is the first step of the process of the measurement of individual health status.

Our work can be potentially used in two ways: 1) more comprehensive and objective measurement of an individual’s health status; 2) development of a health index for an individual. Our results can be used to prioritize the health indicators, e.g., distinguish between a core and a secondary set, and can be referenced by system developers for EHR, personal health records (PHR), or other data capture applications. Our results can contribute to the development of a health index used to stratify healthy research participants to make them more comparable. This is analogous to the Charlson Comorbidity Index[17] or propensity scores[18] that are broadly used in data analytics in clinical epidemiology, both of which, however, are disease-oriented. Although the health indicators reported here are not in a formula format, this is a direction for future research. Our results set the foundation for further weighting, prioritizing, and validating the health indicators via additional data resources. In addition, the measurements can be used in tracking health, measuring the outcomes of preventive services, or aggregating data to examine community health. For health indicators embedded within an EHR or PHR, although having more data points is better, considering clinician burnout[19] in using technology, we need to be more mindful of introducing more specific data capture requirements or expectations to clinical users. In this regard, prioritizing health indicators is a necessary step.

The adjusted clinical group (ACG) system, developed and maintained by Johns Hopkins University over the past 30 years, is a global tool used in population health analytics[20]. The system is rooted in chronic conditions and comorbidities. The goal of the system is fundamentally different from ours, which is to measure individual health (not disease) status more accurately. The Committee on Quality Measures for the Healthy People Leading Health Indicators[3] focuses more on quality measures; they aim to align the measurements with an assessment, improvement, and accountability framework. The focus, however, is on monitoring and reporting at the population level, not necessarily individual health[3].

There are other health-related surveys used broadly worldwide. For example, the SF-36, developed by the RAND Corporation[21], is used to measure life quality and health outcomes. Compared to the SF-12[22], our health indicators provide a more comprehensive measurement, one that goes beyond physical and mental health. PHQ-9[23] is a validated tool to measure depression severity. In our work, however, we are looking for more objective indicators to measure an individual’s health status, both physical and mental.

Our health indicators have fairly good coverage. The Committee on the Recommended Social and Behavioral Domains and Measures for Electronic Health Records of IOM identified measures across the individual and neighborhood levels that involve sociodemographic, psychological, and behavioral data[4, 5]. Among the 17 domains identified by the committee[4, 5], 10 were included in our 29 health indicators. Healthy People 2030[24] proposed 22 leading health indicators for different age groups, of which 16 are included in our health indicators.

### Limitations of the current study

The main limitation of our study is that it is the first step in the determination of the importance of health indicators, and, notably, the results, which are based on public perspectives, are subjective. Further validation of our results via additional objective measures (e.g., healthcare expenditure by disease category[25], burden of illness estimates for specific disease categories[26], etc.) is needed to support our findings.

Another limitation concerns the survey respondents. Females comprised the majority of survey respondents, at 72.1%, 77.7%, and 69% from Ohio University, ResearchMatch, and Clemson University, respectively. We noticed a similar phenomenon in other studies conducted via ReserchMatch. On the one hand, we are happy about the large sample size; on the other hand, the responses may reflect the perspectives of well-educated females more than those of other groups. For example, the survey respondents have an educational level of college and beyond are 54.6%, 82.2%, and 74% among Ohio University, ResearchMatch, and Clemson University respondents.

### Future research

We can foresee several potential directions to continue the project. Our main goal is to validate the results that we obtained via the three surveys we conducted, which can be accomplished in several ways. Because our main goal is to measure individual health status accurately over time, longitudinal data would be ideal. One source is a citizen science project initiated by the National Institutes of Health: all of us[27]; another source is the Biobank initiated in the United Kingdom[28], and the most ideal source is well-documented longitudinal data of a group of individuals that include not only their EHR data but also other data that overlap with our health indicators. Such ideal data sources can provide us with opportunities to examine the corresponding health indicators and to validate the importance of health indicators via EHR records and additional health-related data. In this way, public perspectives will be considered along with more concrete evidence that can ensure more confidence in prioritizing health indicators and using them for various purposes.

## Conclusion

Health indicators are critical for the measurement of individual health status and the determination of preventive services and their outcomes. Obtaining public perspectives on the health indicators is the first step to prioritize these health indicators. The top five rated health indicators are drug and substance abuse, smoking and tobacco use, alcohol abuse, major depression, diet and nutrition. Our respondents, however, had heterogeneous views on the top- and bottom-rated health indicators. The middle 13 health indicators were rated more homogeneously among all the respondents. We separated these 13 health indicators into seven groups or clusters, which provide evidence to further prioritize these 13 health indicators.

## Data Availability

All data produced in the present study are available upon reasonable request to the authors

## Acknowledgments

We want to thank all the respondents who answered the surveys. Without their input, our study would not have been possible.

## Data statement

Data sets and statistical analysis codes used in this manuscript are available via the corresponding author per request.

## Ethics statement

The study was approved by the Institutional Review Boards at Ohio University (17-X-142) and Clemson University (IRB2019-441).

## Author contributions

Conceptualization and design: Xia Jing, Temiloluwa Sokoya, Francisca Lekey, Sebastian Diaz

Acquisition, analysis, or interpretation of data: Xia Jing, Temiloluwa Sokoya, Yuchun Zhou, Sebastian Diaz, Timothy Law, Lina Himawan, Lu Shi, Ronald W. Gimbel

Drafting of the manuscript: Temiloluwa Sokoya, Yuchun Zhou, Xia Jing

Critical revision of the manuscript: Xia Jing, Temiloluwa Sokoya, Yuchun Zhou, Sebastian Diaz, Timothy Law, Lina Himawan, Lu Shi, Ronald W. Gimbel

Statistical analysis: Temiloluwa Sokoya, Yuchun Zhou, Sebastian Diaz, Lina Himawan, Xia Jing

Obtaining funding: Xia Jing

Supervision: Xia Jing, Ronald W. Gimbel

## APPENDICES

**Appendix A**. The online survey instrument

**Appendix B**. Definitions of the health indicators

**Appendix C**. Records from three datasets: before and after data cleaning

**Appendix D**. Codebook

**Appendix E**. Demographic descriptive statistics for all respondents

**Appendix F**. Total of 9 indicators with homogenous variance

**Appendix G**. Total of 16 indicators with heterogeneous variance

**Appendix H**. Total of 16 indicators with significant mean differences post hoc results for the three samples

