## Appendix A for "Health indicators as a measure of individual health status: public perspectives"

### Health indicators to measure an individual's health status: a public perspective survey

Start of Block: Survey consent

Consent Form

Q1.1 This survey is conducted for research purposes through Clemson University. The long-term goal is to set up the foundation to measure preventive services more consistently, accurately, and conveniently. All the data will be collected, analyzed, and reported anonymously. Please contact Xia Jing via *,* Ron Gimbel via, or Lu Shi via for any further questions, comments or suggestions. The estimated survey completion time is between 10-15 minutes. The full-text Exempt adult consent form is provided. By completing this survey I agree that (please ***click all*** options to continue):

- ☐ The respondent is 18 years and older (1)
- ☐ The completion is voluntary (2)
- ☐ The completion and return of the survey implies consent to use the anonymous data for research purposes (4)

End of Block: Survey consent

Start of Block: Respondent's demographic information

Respondent's Demographic Information

Q2.1
Which of the following best describes your age group?

| | | 36-45 (2) | 46-55 (3) | 56-65 (4) | >65 (5) |
|---|---|---|---|---|---|
| Age range (1) | ○ | ○ | ○ | ○ | ○ |

Q2.2 To which gender identity do you most identify?

| | Male (1) | Female (2) | Transgender (3) | Prefer not to answer (4) |
|---|---|---|---|---|
| Gender (1) | ○ | ○ | ○ | ○ |

Q2.3 Which professional group do you belong to?

○ Healthcare providers (1)

○ Public health professional (2)

○ Researcher who uses health indicator data (3)

○ Other researchers (4)

○ Others (Specify) (5) ________________________________________________

Q2.4 Please select your highest educational qualification.

| | High school (1) | Associate degree (2) | College degree (3) | Master degree (4) | Doctoral degree (5) |
|---|---|---|---|---|---|
| Educational level (1) | ○ | ○ | ○ | ○ | ○ |

Q2.5 To which racial background do you primarily identify?

| | White American (1) | African American (2) | Hispanic and Latino American (3) | Asian American (4) | Native American (Including Alaska native, native Hawaiian and other Pacific Islander (5) | Two or more races (6) | Other (7) |
|---|---|---|---|---|---|---|---|
| Race Groups (1) | ○ | ○ | ○ | ○ | ○ | ○ | ○ |

Q2.6 Which country do you live in?

○ United States (1)

○ Other(specify) (2) ________________________________________________

*Display This Question:*

*If Which country do you live in? = United States*

Q2.7 Which state do you live in the U.S?

| | |
|---|---|
| State (1) | ▼ AL (1) ... WY (50) |

Page Break

End of Block: Respondent's demographic information

Start of Block: Block 2

Q3.1 What can be used to measure an individual's general health status (i.e., refer to a person's overall physical, mental, and social well-being, free from illness or injury)? Your view counts!
From the scale of **0** (**not at all important**) to **10** (**extremely important**) how important do you think each health indicator can be used to measure an individual's health status? Please move the slide bar to answer for each indicator.

*

Q3.2 **Health risks and behavior Indicators**

| | 0 1 2 3 4 5 6 7 8 9 10 |
|---|---|
| Alcohol abuse (1) | |
| Body Mass Index (BMI) (2) | |
| Diet and nutrition (3) | |
| Drug or substance abuse (4) | |
| Family history of cancer (5) | |
| Physical inactivity (6) | |
| Smoking tobacco use (7) | |
| Sun protection (8) | |

Q3.3 **Healthcare**

| | 0 | 1 | 2 | 3 | 4 | 5 | 6 | 7 | 8 | 9 | 10 |
|---|---|---|---|---|---|---|---|---|---|---|---|
| Immunization/Vaccination (1) | | | | | | | | | | | |
| Insurance coverage (2) | | | | | | | | | | | |
| Personal care needs (3) | | | | | | | | | | | |

Q3.4 **Healthcare provider supply**

| | 0 | 1 | 2 | 3 | 4 | 5 | 6 | 7 | 8 | 9 | 10 |
|---|---|---|---|---|---|---|---|---|---|---|---|
| Cancer screening detection (1) | | | | | | | | | | | |
| Hypertension screening (2) | | | | | | | | | | | |
| HIV testing (3) | | | | | | | | | | | |

Q3.5 **Health related quality of life**

| | 0 | 1 | 2 | 3 | 4 | 5 | 6 | 7 | 8 | 9 | 10 |
|---|---|---|---|---|---|---|---|---|---|---|---|
| Self-rated health status (1) | | | | | | | | | | | |

Q3.6 **Blood tests in physical exams**

| | 0 | 1 | 2 | 3 | 4 | 5 | 6 | 7 | 8 | 9 | 10 |
|---|---|---|---|---|---|---|---|---|---|---|---|
| Blood sugar level (2) | | | | | | | | | | | |
| Blood triglycerides (3) | | | | | | | | | | | |
| HDL cholesterol (4) | | | | | | | | | | | |
| LDL cholesterol (5) | | | | | | | | | | | |
| Total cholesterol (6) | | | | | | | | | | | |

Q3.7 **Education**

| | 0 | 1 | 2 | 3 | 4 | 5 | 6 | 7 | 8 | 9 | 10 |
|---|---|---|---|---|---|---|---|---|---|---|---|
| High school diploma as a health indicator (1) | | | | | | | | | | | |

Q3.8 This is a verification question: please select the obvious option that does not belong to the same category as other options.

- ○ Apple (1)
- ○ Orange (2)
- ○ Beef (3)
- ○ Banana (4)
- ○ Blueberry (5)

---

Q3.9
**Other Health Indicators**

| | 0 | 1 | 2 | 3 | 4 | 5 | 6 | 7 | 8 | 9 | 10 |
|---|---|---|---|---|---|---|---|---|---|---|---|
| Air quality index > 100 (1) | | | | | | | | | | | |
| Dentist supply (2) | | | | | | | | | | | |
| Engaged people (people's attention or efforts are occupied) (3) | | | | | | | | | | | |
| Health literacy rate (4) | | | | | | | | | | | |
| Major depression (5) | | | | | | | | | | | |
| Having a sense of purpose in one's life (6) | | | | | | | | | | | |
| Race and ethnicity (7) | | | | | | | | | | | |
| Unemployed individual (8) | | | | | | | | | | | |

Q24 Your overall selection of the most important **top three** health indicators: Please ONLY select the top three. If you would like to specify other indicators, please select "Other indicator" and specify it.

- ☐ Air quality index >100 (1)
- ☐ Alcohol abuse (2)
- ☐ Blood sugar level (3)
- ☐ Blood triglycerides (4)
- ☐ Body mass index (BMI) (5)
- ☐ Cancer screening detection (6)
- ☐ Dentist supply (7)
- ☐ Diet and nutrition (8)
- ☐ Drug or substance abuse (9)
- ☐ Engaged people (people's attention or efforts are occupied) (10)
- ☐ Family history of cancer (11)
- ☐ HDL cholesterol (12)
- ☐ Health literacy rate (13)
- ☐ High school diploma (14)
- ☐ HIV testing (15)
- ☐ Hypertension screening (16)
- ☐ Immunization/Vaccination (17)

- [ ] Insurance coverage (18)
- [ ] LDL cholesterol (19)
- [ ] Major depression (20)
- [ ] Personal care needs (21)
- [ ] Physical inactivity (22)
- [ ] Having a sense of purpose in one's life (23)
- [ ] Race and ethnicity (24)
- [ ] Self-rated health status (25)
- [ ] Smoking, tobacco use (26)
- [ ] Sun protection (27)
- [ ] Total cholesterol (28)
- [ ] Unemployed individual (29)
- [ ] Other indicator(please specify) (30) ________________

Q3.10 Are there any other important primary health indicators that should be included? Please specify:

________________________________

________________________________

________________________________

________________________________

End of Block: Block 2

Start of Block: Your perspective about healthy

Q4.1 Which of the following statement(s) can be counted as healthy in your view?Please ***click all*** your answers.

- ▢ A person has no diagnosed diseases/conditions (1)
- ▢ A person has clear diagnosed disease/condition, however the diseases/conditions are well controlled and well managed (2)
- ▢ A person has good mental (i.e., emotional, psychological and social well-being) and physical (i.e., perform daily activities without problems) statuses regardless of their diagnoses (3)
- ▢ A person thinks he/she is in good health regardless of symptoms or diagnoses (4)

Q4.2 Please add any additional thoughts about the definition of healthy.

________________________________________________________________

________________________________________________________________

________________________________________________________________

________________________________________________________________

________________________________________________________________

Q4.3 Do you have other comments, suggestions or perspectives you would like to add to this topic?

________________________________________________________________

________________________________________________________________

End of Block: Your perspective about healthy
