## Appendix B for "Health indicators as a measure of individual health status: public perspectives"

| HI definitions provided via surveys | |
|---|---|
| **HI** | **Definition** |
| Alcohol abuse | Alcohol misuse describes alcohol consumption that puts individuals at increased risk for adverse health and social consequences. It is defined as excess daily consumption (more than 4 drinks per day for men or more than 3 drinks per day for women), or excess total consumption (more than 14 drinks per week for men or more than 7 drinks per week for women), or both. |
| Blood sugar level | The fasting glucose level in blood. |
| Blood triglycerides | Triglycerides are a type of fat found in your blood that your body uses for energy. The combination of high levels of triglycerides with low HDL cholesterol or high LDL cholesterol can increase your risk for heart attack and stroke. |
| BMI | 1.Body Mass Index (BMI) is a person's weight in kilograms divided by the square of height in meters.<br>2.Class 1: BMI of 30 to < 35<br>3.Class 2: BMI of 35 to < 40<br>4.Class 3: BMI of 40 or higher<br><br>1.If your BMI is less than 18.5, it falls within the underweight range.<br>2.If your BMI is 18.5 to <25, it falls within the normal.<br>3.If your BMI is 25.0 to <30, it falls within the overweight range. |
| Cancer screening detection | e.g., cervical cancer screening (pap), breast cancer screening (mammogram), Colorectal cancer screening. |
| HDL cholesterol | HDL cholesterol absorbs cholesterol and carries it back to the liver, which flushes it from the body. HDL is known as “good” cholesterol because having high levels can reduce the risk for heart disease and stroke. |
| Health literacy rate | Health literacy refers, broadly, to the ability of individuals to “gain access to, understand and use information in ways which promote and maintain good health”1 for themselves, their families and their communities. |
| HIV testing | A medical condition where the immune system cannot function properly and protect the body from disease. As a result, the body cannot defend itself against infections (like pneumonia). AIDS is caused by the Human Immunodeficiency Virus (HIV). This virus is spread through direct contact with the blood and body fluids of an infected individual. High risk activities include unprotected sexual intercourse and intravenous drug use (sharing needles). |
| Hypertension screening | Percentage of patients 18-85 years of age who had a diagnosis of hypertension and whose blood pressure was adequately controlled (<140/90 mmHg) during the measurement period. |
| Immunization vaccination | Immunization is the process whereby a person is made immune or resistant to an infectious disease, typically by the administration of a vaccine. Vaccines stimulate the body’s own immune system to protect the person against subsequent infection or disease. |

| | |
|---|---|
| Insurance coverage | The Insured category includes persons who have private health insurance, Medicaid, military coverage, Medicare, Children’s Health Insurance Program (CHIP), state-sponsored or other government-sponsored health plan, Medicare (disability), or military plan (TRICARE, Veterans Administration (VA) and CHAMP-VA). A person was not considered to be insured if he or she had only Indian Health Service coverage or only had a private plan that paid for one type of service such as accidents or dental care. |
| LDL cholesterol | LDL cholesterol makes up the majority of the body’s cholesterol. LDL is known as “bad” cholesterol because having high levels can lead to plaque buildup in your arteries and result in heart disease and stroke. |
| Personal care needs | Personal care needs, or activities of daily living, include eating, bathing, dressing, and getting around inside the person’s home. |
| Physical inactivity | Any bodily movement produced by the contraction of skeletal muscle that increases energy expenditure above a basal level. In these Guidelines, physical activity generally refers to the subset of physical activity that enhances heal |
| Self rated health status | The assessment of the individual level of “awareness” regarding their health status. Demographic characteristics, health care coverage, leisure-time physical activity, and body mass index may predict self-rated health status. |
| Smoking, tobacco use | Tobacco products are products made entirely or partly of leaf tobacco as raw material, which are intended to be smoked, sucked, chewed or snuffed. All contain the highly addictive psychoactive ingredient, nicotine |
| Substance abuse | Substance abuse refers to the harmful or hazardous use of psychoactive substances, including alcohol and illicit drugs. |
| Total cholesterol | Cholesterol is a fat-like substance, found in the bloodstream as well as in bodily organs and nerve fibres. |
