## Appendix C for "Health indicators as a measure of individual health status: public perspectives"

**Appendix C**. Records from three datasets: before and after data cleaning

| | **Agreed and valid (based on validation question)** | **Incomplete records** | **Complete records** | **Invalid records (less than 3 mins)** | **Complete and valid records for analysis (after cleaning)** |
|---|---|---|---|---|---|
| Ohio University dataset | 389 | 4 | 385 | 23 | 362 |
| Clemson dataset | 182 | 80 | 102 | 5 | 97 |
| ResearchMatch | 694 | 9 | 685 | 0 | 685 |
