## Appendix D for "Health indicators as a measure of individual health status: public perspectives"

**Appendix D**. Codebook

| **Variables** | **Range** | **Notes for each code** |
|---|---|---|
| Age | [1-5] | 1: < = 35<br>2: 36-45<br>3: 46-55<br>4: 56-65<br>5: > 65 |
| Gender | [1-4] | 1: Female<br>2: Male<br>3: Transsexual<br>4: Prefer not to answer |
| Professional group | [1-5] | 1: Healthcare providers<br>2: Public health professional<br>3: Researcher who uses health indicator data<br>4: Other researchers<br>5: Other professional groups (specify) |
| Educational qualification | [1-5] | 1: High school<br>2: Associate's degree<br>3: Bachelor's degree<br>4: Master's degree<br>5: Doctoral degree |
| Race | [1-7] | 1: White American<br>2: African American<br>3: Hispanic and Latino American<br>4: Asian American<br>5: Native American (including Alaska |

| | | |
|---|---|---|
| | | native, native Hawaiian, and other Pacific Islander)<br>6: Two or more races<br>7: Other |
| Sample | [1-3] | 1: ResearchMatch<br>2: OU<br>3: Clemson |
