## Appendix E for "Health indicators as a measure of individual health status: public perspectives"

**Appendix E.** Demographic descriptive statistics for all respondents

| | | Frequency (n) | Percent (%) |
|---|---|---|---|
| Source | ResearchMatch | 694 | 60.2 |
| | Ohio University | 362 | 31.4 |
| | Clemson University | 97 | 8.4 |
| | Total | 1153 | 100 |
| Age Group | < = 35 | 464 | 40.2 |
| | 36-45 | 163 | 14.1 |
| | 46-55 | 152 | 13.2 |
| | 56-65 | 201 | 17.4 |
| | > 65 | 173 | 15 |
| | Total | 1153 | 100 |
| Gender | Female | 862 | 74.8 |
| | Male | 273 | 23.7 |
| | Transgender | 10 | 0.9 |
| | Prefer not to answer | 8 | 0.7 |
| | Total | 1153 | 100 |
| Professional Group | Healthcare providers | 232 | 20.1 |
| | Public health professional | 71 | 6.2 |
| | Researcher who uses health indicator data | 78 | 6.8 |
| | Other researchers | 204 | 17.7 |
| | Other professional groups (Specify) | 568 | 49.3 |
| | Total | 1153 | 100 |
| Educational qualification | High school | 218 | 18.9 |
| | Associate degree | 95 | 8.2 |
| | College degree | 367 | 31.8 |
| | Master degree | 327 | 28.4 |
| | Doctoral degree | 146 | 12.7 |
| | Total | 1153 | 100 |
| Race | White American | 1006 | 87.3 |
| | African American | 38 | 3.3 |
| | Hispanic and Latino American | 25 | 2.2 |
| | Asian American | 19 | 1.6 |
| | Native American (Including Alaska native, native Hawaiian and other Pacific Islander) | 5 | 0.4 |
| | Two or more races | 23 | 2 |

| | | | |
|---|---|---|---|
| | Other | 37 | 3.2 |
| | Total | 1153 | 100 |
