## Appendix F for "Health indicators as a measure of individual health status: public perspectives"

**Appendix F**. Total of 9 indicators with homogenous variance

| **Health indicator** | **Levene test Sig.** |
|---|---|
| Blood sugar level | 0.19 |
| HDL cholesterol | 0.22 |
| LDL cholesterol | 0.21 |
| Total cholesterol | 0.83 |
| Immunization/vaccination | 0.12 |
| Insurance coverage | 0.07 |
| Cancer screening detection | 0.05 |
| Air quality index > 100 | 0.14 |
| Self-rated health status | 0.34 |
