## Appendix G for "Health indicators as a measure of individual health status: public perspectives"

**Appendix G**. Total of 16 indicators with heterogeneous variance

| **Health indicator** | **LevenetTest Sig.** |
|---|---|
| Blood triglycerides | 0.05 |
| Alcohol abuse | 0.004 |
| Body mass index (BMI) | 0.002 |
| Diet and nutrition | 0.001 |
| Drug or substance abuse | 0.00 |
| Family history of cancer | 0.05 |
| Physical inactivity | 0.00 |
| Smoking, tobacco use | 0.00 |
| Sun protection | 0.00 |
| Personal care needs | 0.05 |
| Hypertension screening | 0.00 |
| HIV testing | 0.03 |
| High school diploma as a health indicator | 0.00 |
| Supply of dentists | 0.03 |
| Engagement in life | 0.00 |
| Health literacy rate | 0.03 |
| Major depression | 0.01 |
| Having a sense of purpose in one’s life | 0.00 |
| Race and ethnicity | 0.04 |
| Unemployment | 0.01 |
