## Appendix H for "Health indicators as a measure of individual health status: public perspectives"

**Appendix H.** Total of 16 indicators with significant mean differences post hoc results for the three samples

| **Health indicator** | | | **Post hoc Sig.** |
|---|---|---|---|
| | ResearchMatch | Clemson | 0.38 |
| | OU | ResearchMatch | 0.00 |
| Alcohol abuse | Clemson | OU | 0.052 |
| | ResearchMatch | Clemson | 0.70 |
| | OU | ResearchMatch | 0.00 |
| Body mass index (BMI) | Clemson | OU | 0.01 |
| | ResearchMatch | Clemson | 0.16 |
| | OU | ResearchMatch | 0.18 |
| Diet and nutrition | Clemson | OU | 0.02 |
| | ResearchMatch | Clemson | 0.10 |
| | OU | ResearchMatch | 0.00 |
| Drug or substance abuse | Clemson | OU | 0.50 |
| | ResearchMatch | Clemson | 0.01 |
| | OU | ResearchMatch | 0.00 |
| Family history of cancer | Clemson | OU | 0.88 |
| | ResearchMatch | Clemson | 0.54 |
| | OU | ResearchMatch | 0.00 |
| Physical inactivity | Clemson | OU | 0.45 |
| | ResearchMatch | Clemson | 0.004 |
| | OU | ResearchMatch | 0.00 |
| Smoking, tobacco use | Clemson | OU | 0.74 |

| | | | |
|---|---|---|---|
| | ResearchMatch | Clemson | 0.00 |
| | OU | ResearchMatch | 0.00 |
| Sun protection | Clemson | OU | 0.73 |
| | ResearchMatch | Clemson | 0.02 |
| | OU | ResearchMatch | 0.12 |
| Insurance coverage | Clemson | OU | 0.35 |
| Hypertension screening | ResearchMatch | Clemson | 0.75 |
| | OU | ResearchMatch | 0.01 |
| | Clemson | OU | 0.56 |
| Engagement in life | ResearchMatch | Clemson | 1.00 |
| | OU | ResearchMatch | 0.00 |
| | Clemson | OU | 0.00 |
| Major depression | ResearchMatch | Clemson | 0.94 |
| | OU | ResearchMatch | 0.02 |
| | Clemson | OU | 0.46 |
| Having a sense of purpose in one’s life | ResearchMatch | Clemson | 0.47 |
| | OU | ResearchMatch | 0.00 |
| | Clemson | OU | 0.00 |
| Race and ethnicity | ResearchMatch | Clemson | 0.66 |
| | OU | ResearchMatch | 0.00 |
| | Clemson | OU | 0.07 |
| High school diploma | ResearchMatch | OU | 0.00 |

| | | | |
|---|---|---|---|
| | OU | Clemson | 0.31 |
| | Clemson | ResearchMatch | 0.20 |
| Unemployment | ResearchMatch | Clemson | 0.90 |
| | OU | ResearchMatch | 0.00 |
| | Clemson | OU | 0.06 |

IV = the three samples, DV = Individual survey data.
